# Altered subcortical emotional salience processing differentiates Parkinson’s patients with and without psychotic symptoms

**DOI:** 10.1101/19010512

**Authors:** F. Knolle, S. Garofalo, R. Viviani, A. Justicia, A.O. Ermakova, H. Blank, G.B. Williams, G. Arrondo, P. Ramachandra, C. Tudor-Sfetea, N. Bunzeck, E. Duezel, T.W. Robbins, R.A. Barker, G.K. Murray

## Abstract

**Background:** Current research does not provide a clear explanation for why some patients with Parkinson’s Disease (PD) develop psychotic symptoms. In the field of schizophrenia research the ‘aberrant salience hypothesis’ of psychosis has been influential. According to the theory, dopaminergic dysregulation leads to the inappropriate attribution of salience to otherwise irrelevant or non-informative stimuli, allowing for the formation of hallucinations and delusions. This theory has not yet been extensively investigated in the context of psychosis in PD.

**Methods:** We investigated salience processing in 14 PD patients with a history of psychotic symptoms, 23 PD patients without psychotic symptoms and 19 healthy controls. All patients received dopaminergic medication. We examined emotional salience using a visual oddball fMRI paradigm (Bunzeck and Düzel, 2006) that previously has been used to investigate early stages of schizophrenia spectrum psychosis, controlling for resting cerebral blood flow as assessed with arterial spin labelling fMRI.

**Results:** We found significant differences between patient groups in brain responses to emotional salience. PD patients with psychotic symptoms revealed senhanced brain responses in the striatum, the hippocampus and the amygdala compared to PD patients without psychotic symptoms. PD patients with psychotic symptoms also showed significant correlations between the levels of dopaminergic drugs they were taking and BOLD signalling, as well as psychotic symptom scores. Furthermore, our data provide a first indication for dysfunctional top-down processes, measured in a ‘jumping to conclusions’ bias.

**Conclusion:** Our study suggests that enhanced signalling in the striatum, hippocampus and amygdala together with deficient top-down cognitive regulations is associated with the development of psychotic symptoms in PD which is similar to that proposed in the ‘aberrant salience hypothesis’ of psychosis in schizophrenia.

## Introduction

Parkinson’s disease (PD) patients frequently suffer from psychotic symptoms, especially as the disease advances, which most commonly takes the form of visual hallucinations, delusions and illusions (Aarsland *et al*, 1999). With disease progression psychotic symptoms may shift to other modalities such as the auditory domain, comprising auditory hallucinations of incomprehensible voices (Inzelberg *et al*, 1998) or non-verbal sounds (Fenelon, 2000). PD psychosis characterises a spectrum of such psychotic symptoms that occur throughout the course of the disease, but especially in those with longer disease duration, giving an overall prevalence of 26% (Mack *et al*, 2012) (Forsaa *et al*, 2010; Gibson *et al*, 2013). Its development is associated with increased risk for mortality and nursing home placement as well as lower overall global functioning and well-being (Ffytche *et al*, 2017).

Current research suggests that PD psychosis combines a set of symptoms with a specific pathophysiology (comprehensive review (Ffytche *et al*, 2017)), rather than a single mechanistic dysfunction. Risk and modulatory factors include genetics, the use of dopamine-based antiparkinsonian drugs, and disease-specific factors such as cognitive impairment, dementia, duration and severity, depression, sleep disorders, and age and the presence of intercurrent infections or illnesses (Fénelon and Alves, 2010; Friedman, 2010). Although there are clear differences between the primary psychiatric disorder of (schizophrenia spectrum) psychosis and PD psychosis, a disturbed dopaminergic system is a unifying element in both diseases, possibly contributing to the occurrence of psychotic symptoms in both disorders (Carter and Ffytche, 2015; Garofalo *et al*, 2017). PD psychosis is particularly interesting as it is commonly found as a comorbidity in PD patients but does not affect all.

A dysfunctional dopaminergic signal, perhaps in the mesolimbic regions, is argued to be associated with the inappropriate attribution of salience to otherwise irrelevant or non-informative stimuli, allowing for the formation of hallucinations and delusions; this theory has been termed the ‘aberrant salience hypothesis’ of psychosis (Heinz, 2002; Kapur, 2003; Roiser *et al*, 2013). Some models propose that within the hippocampal-striatal-midbrain circuits, hippocampal dysfunction leads to an enhanced subcortical dopaminergic signalling through descending projections to the dorsal striatum (Lisman and Grace, 2005; Lodge and Grace, 2007). Supporting the involvement of these circuits, a recent study investigating novelty salience processing reported increased connectivity of hippocampal to striatal and midbrain regions, but decreased connectivity between the striatum and the midbrain in subjects at high risk of developing psychosis (Modinos *et al*, 2019). Furthermore, our previous work in first-episode psychosis patients, using the same salience paradigm as Modinos et al, showed reduced midbrain, striatal and occipital activation while processing novelty and negative emotional salient stimuli (Knolle *et al*, 2018). In Parkinson’s disease research, the investigation of salience processing has received comparably little attention. One study showed that the use of a dopamine agonist (pramipexole or ropinirole) in young, medication-naïve PD patients led to an increase in aberrant motivational salience by facilitating arbitrary and illusory associations between stimuli and rewards, and by faster reaction times to task-irrelevant stimuli (Nagy *et al*, 2012). Unmedicated patients in that latter study did not suffer from psychotic symptoms, but had increased scores on the O-LIFE unusual experience score after treatment with dopaminergic agents (Nagy *et al*, 2012). Furthermore, another study (Mannan *et al*, 2008) suggested impaired salience processing in PD: in an eye-gaze experiment, patients showed an impaired ability to detect a salient stimulus in a visual search task. No psychotic symptoms were reported for the patients in that latter study. In our previous work (Garofalo *et al*, 2017), we investigated reward processing, a form of motivation salience, in PD patients with and without psychotic symptoms and in controls. PD patients with psychotic symptoms showed very similar patterns of reduced activation (including in the striatum and cingulate cortex) as reported in primary psychosis individuals (Ermakova *et al*, 2018; Murray *et al*, 2008).

In this new study we report here, we sought to explore whether the ‘aberrant salience hypothesis’ of psychosis can be applied to PD psychosis and thus provide an explanation as to how psychotic symptoms arise in PD patients. To our knowledge, the current study is the first to investigate brain responses to non-motivational salient visual stimuli in PD psychosis. We specifically investigated salience processing in PD patients with psychotic symptoms, in PD patients without psychotic symptoms and in healthy controls. We used an fMRI salience paradigm (Bunzeck and Düzel, 2006) that previously has shown significantly altered midbrain, striatal, hippocampal and amygdala activations and connectivity in early stages of “psychiatric” psychosis in young adults (Knolle *et al*, 2018; Modinos *et al*, 2019). Based on the literature and our previous findings, we hypothesised, first, that PD psychosis patients would demonstrate altered brain responses in the substantia nigra, the striatum, the hippocampus and the amygdala compared to healthy controls and, second, that PD patients without psychotic symptoms would show intermediate processing in response to negative emotional salience.

The aberrant salience theory of psychosis has posited that whilst perceptual salience may be misattributed in psychosis, higher-order cognitive processes are invoked to shape abnormal experiences into abnormal beliefs. Whilst our focus in the current study is on brain correlates of emotional salience processing, in a preliminary analysis we also examined whether higher-order (probabilistic) reasoning is affected in PD psychosis.

## Methods

### Subjects

In total, we recruited 26 participants, who had a diagnosis of PD without any psychotic symptoms using established diagnostic criteria; 15 participants with a diagnosis of PD and ongoing or previous psychotic symptoms, and 19 healthy control subjects, without any history of neurological or psychiatric disorder, matched in age, gender, education (see Table 1). We assessed psychotic symptoms in Parkinson’s patients using the Comprehensive Assessment of At Risk Mental States (CAARMS) (Yung *et al*, 2005)-also see our previous work (Garofalo *et al*, 2017) for a detailed description and Table 1). For two participants, one healthy control and one PD patient with psychotic symptoms, the fMRI session had to be aborted during scanning of the relevant task, as both participants felt uncomfortable inside the scanner. Both participants decided not to continue with the scanning and so were excluded from any analysis. Additionally, two PD patients without psychotic symptoms were excluded due to excessive movement artefact in the scanner (see details below). Finally, two outliers were identified during our analysis, one healthy control and one PD patient without psychotic symptoms, who exceeded -/+ two standard deviations from the averaged imaging signal in all regions of interest (ROI). The final sample therefore comprised of 52 participants: 23 PD patients without psychotic symptoms, 17 healthy controls and 14 PD patients with psychotic symptoms.

**Table 1:**
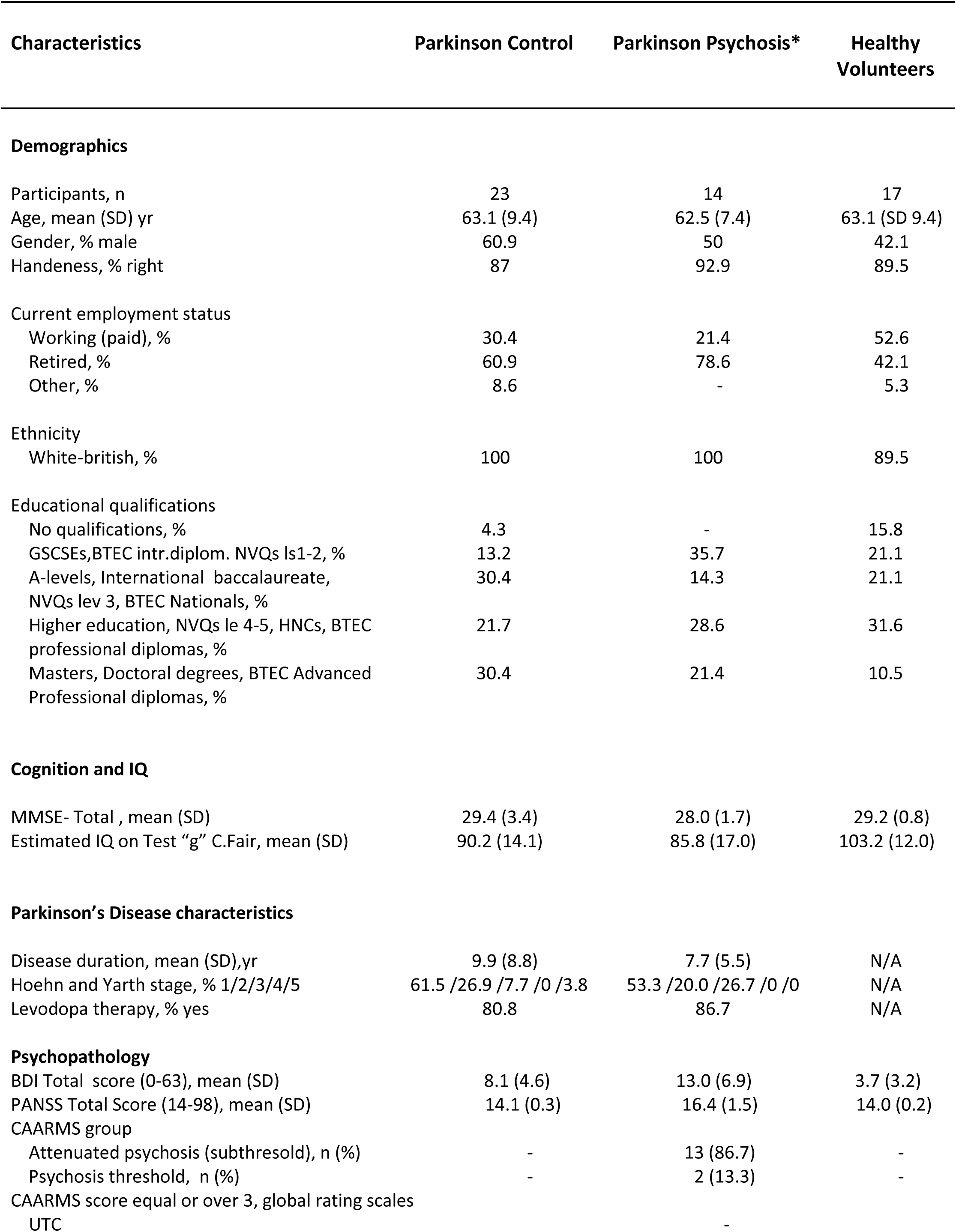
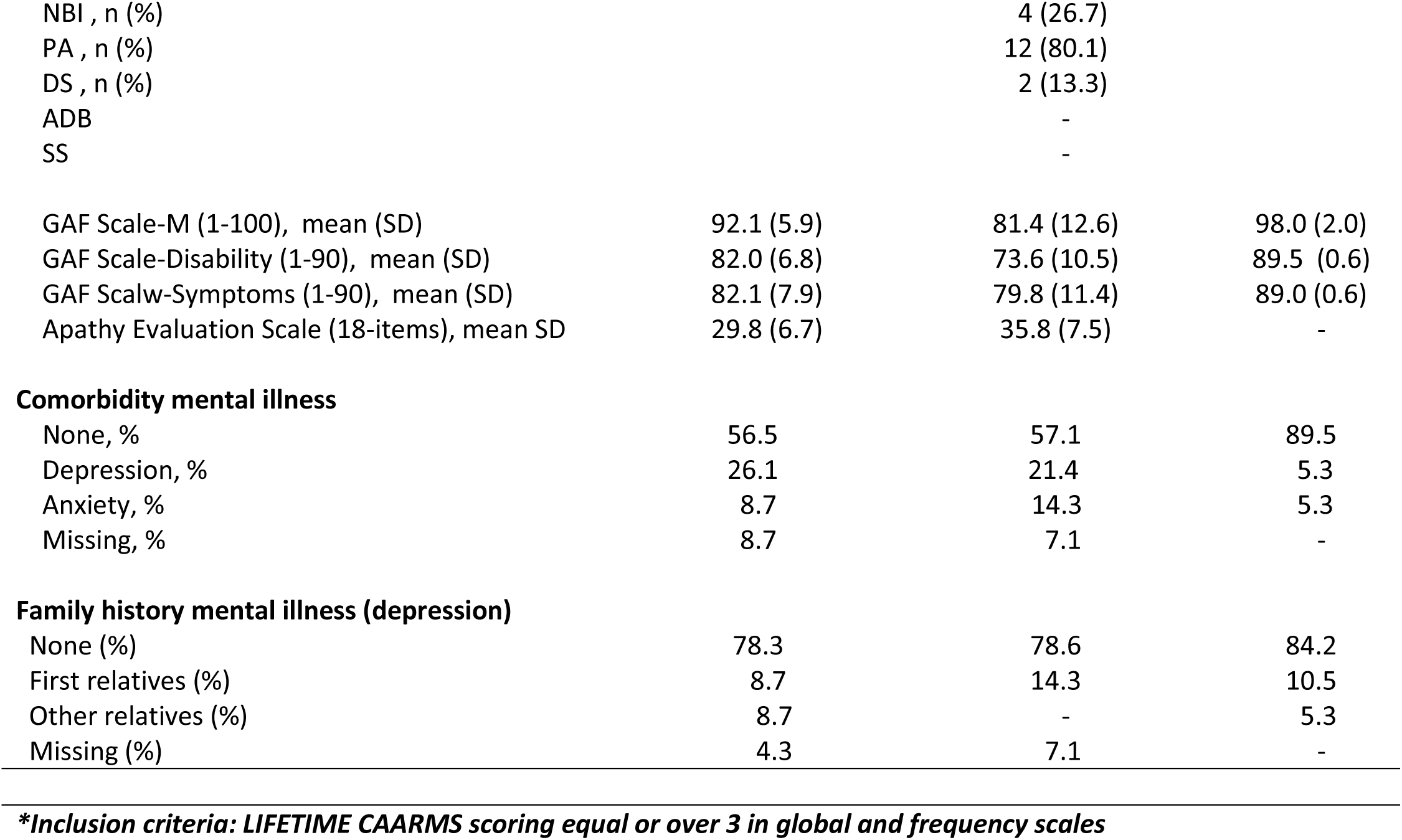
Demographics and pathology of psychiatric aspects of Parkinson’s Disease

Patients were recruited via the PD research clinic at the John van Geest Centre for Brain Repair (VGB). All patients met the Queen Square Brain Bank Criteria for idiopathic PD (Gibb and Lees, 1988). Patients with dementia were excluded (mini-mental state score less than 24). In all cases, the patients anti-PD medication remained unchanged during testing, and was converted to a LED using a standard approach (Tomlinson *et al*, 2010). The two patient groups did not significantly differ in the LED they received (p=.572). Before scanning, each of the participants underwent a general interview and clinical assessment using the Positive and Negative Symptom Scale (PANSS) (Kay *et al*, 1987), and the Global Assessment of Functioning (GAF) (Hall, 1995). The Beck Depression Inventory (BDI) (Beck *et al*, 1996) was used to assess depressive symptoms during the last two weeks and IQ was estimated using the Culture Fair Intelligence Test (Cattell and Cattell, 1973) and cognitive impairment was measured using the mini-mental state examination (Folstein *et al*, 1975).

All subjects had normal or corrected-to-normal visual acuity and were without any contraindications for MRI scanning. At the time of the study, none of the participants were taking antipsychotic medications or had drug or alcohol dependence. The study was approved by the Cambridgeshire 3 National Health Service research ethics committee. All subjects gave written informed consent in accordance with the Declaration of Helsinki.

### Salience oddball task

We used a visual oddball paradigm (Bunzeck *et al*, 2007) in order to investigate negative emotional salience (Figure 1). A detailed description of the paradigm can be found in our previous work (Knolle *et al*, 2018): in summary, participants saw a series of greyscale images of faces and outdoor scenes. 66.6% of these were ‘standard’, neutral images. Then four types of deviating images were randomly intermixed with these; each type occurred with a probability of 8.3%. These deviant events were: stimuli that evoked a negative emotional response (‘emotional oddball’, angry face or image of car crash); neutral stimuli that required a motor response (‘target oddball’); neutral stimuli that presented a novel image every time they appeared (‘novel oddball’); and neutral stimuli of the same face or scene that did not require a motor response or contained negative/positive emotional valence (‘neutral oddball’) (Figure 1). All participants completed 3 blocks of 240 trials each (160 standard trials, and 20 oddball trials each of target, neutral, emotional and novel stimuli), resulting in a total of 720 trials. The task contained 50% faces and 50% outdoor scenes, to avoid category-specific habituation and to make stimulus exploration biologically relevant. Participants were introduced to the target stimulus prior to the experimental session for 4.5s, and they were required to make a simple button press in response using their left or right index finger to each of its subsequent appearances during the experiment within the fMRI-scanner. Participants used their preferred or less affected hand to press the buttons on the button box for the target picture. No motor responses were associated with any of the other stimulus types.

**Figure 1.**
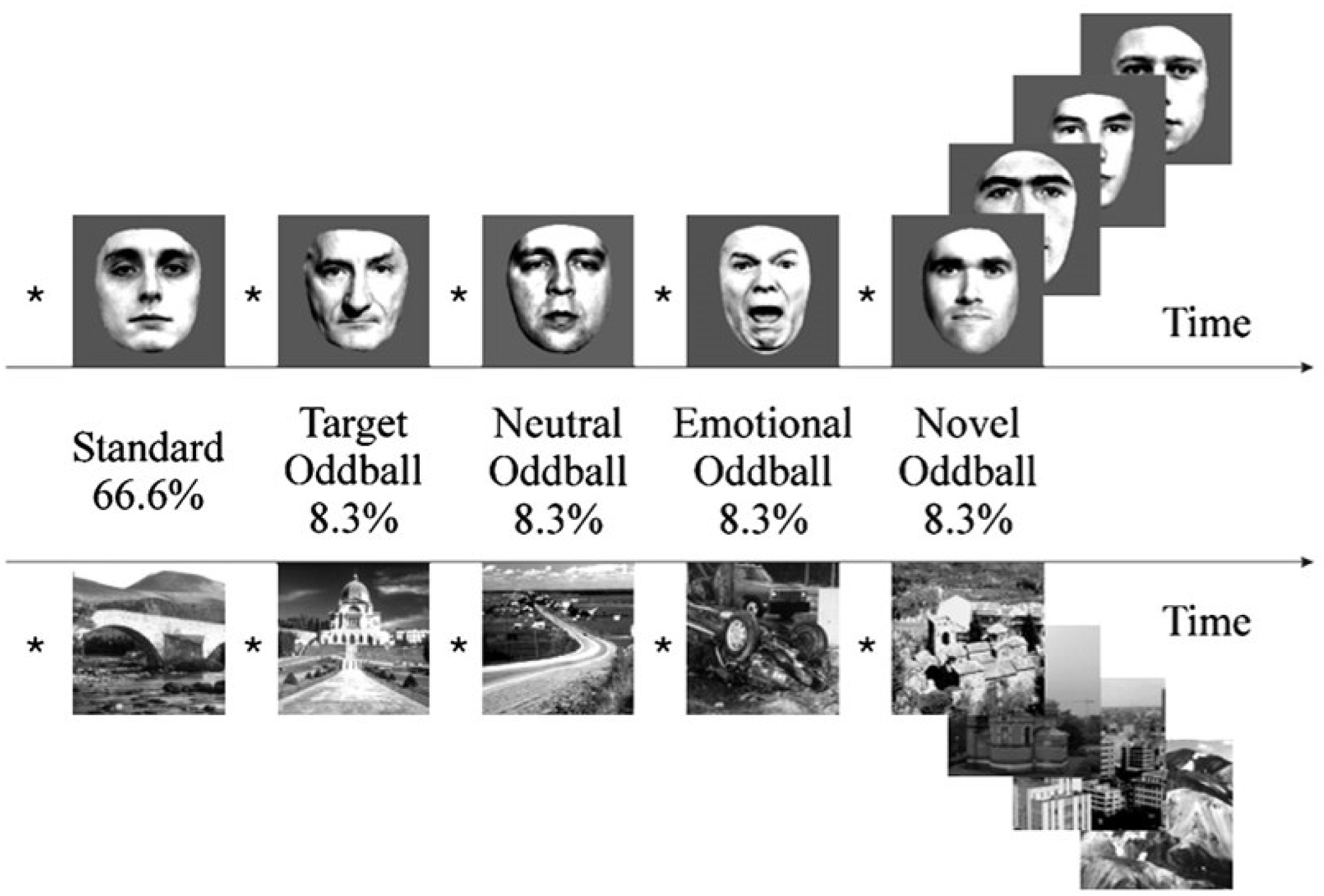
Task design. During this visual oddball paradigm, participants are presented with a random series of greyscale images of faces and outdoor scenes. 66.6% of these were ‘standard’ images. The remaining 33.4% consisted of four types of rare or contextually deviant events, which were randomly intermixed with the standard stimuli; each occurred with a probability of 8.3%. These deviant events were: neutral stimuli that required a motor response (‘target oddball’); stimuli that evoked a negative emotional response (‘emotional oddball’, angry face or image of car crash); novel stimuli (‘novel oddball’, different neutral images that appear only once); and neutral stimuli (‘neutral oddball’, neutral image of face or scene). In the current study, we were only interested in the contrast between negative emotional and neutral oddball stimuli (image from (Bunzeck and Düzel, 2006)).

During the fMRI-experiment, the pictures were presented for 500ms followed by a white fixation cross on a grey background (grey value=127) using an inter-stimulus interval (ISI) of 2.7s. ISI was jittered with ±300ms (uniformly distributed). All of the stimuli were taken from Bunzeck and Düzel (Bunzeck and Düzel, 2006). The scalp hair and ears of faces were removed artificially; the outdoor scenes did not include faces. All pictures were grey scaled and normalised to a mean grey value of 127 (SD 75). The pictures were projected on to the centre of a screen, and the participants watched them through a mirror mounted on the head coil, subtending a visual angle of about 8°.

In the current study, we focussed on negative emotional salience, this contrast was the most robust in terms of generating within and between group brain activations in our previous study in young adults with first-episode psychosis (Knolle *et al*, 2018). We contrasted activation associated with the emotional and neutral oddball. The contrast between these stimuli allowed us to examine brain responses to the pure stimulus negative emotional valence (‘emotional’ vs. ‘neutral’) (Figure 1) given that both images appear at the same frequency. In contrast, the classical oddball effect is sought by looking at the contrast between neutral oddball and the standard stimuli, which is based on frequency differences.

### Additional behavioural testing: probabilistic reasoning

Additionally, some participants completed a ‘jumping to conclusion’ task (Ermakova *et al*, 2019): 12 of 23 PD patients without psychosis, 5 of 15 patients with psychotic symptoms, and 18 of 19 controls subjects. Due to the incomplete nature of the data we report these data in a preliminary analysis in the supplementary materials.

### Behaviour analysis

An analysis of variance (ANOVA) was used to investigate group differences in responses to the target stimuli (i.e. button presses) as well as reaction times. All blocks in which participants missed more than five button presses were excluded. Behavioural data were analysed using SPSS 21 (IBM Corp.).

### Neuroimaging acquisition and analysis protocol

Data were collected using a Siemens Magnetom Trio Tim syngo MR B17 operating at 3 T.

We used a previously described protocol for the acquisition of the functional imaging data (Knolle *et al*, 2018). We acquired gradient-echo echo-planar T2*-weighted images depicting BOLD contrast from 35 non-contiguous oblique axial plane slices of 2mm thickness to minimise signal drop-out in the ventral regions. No images of the whole brain were retrieved; the superior part of the cortex was not imaged. We used the following setup: relaxation time: 1620m; echo time: 30ms; flip angle: 65°; in-plane resolution: 3×3×3mm; matrix size: 64×64; field of view: 192×192mm; and bandwidth: 2442Hz/px. We acquired a total of 437 volumes per participant (35 slices each of 2mm thickness, inter-slice gap 1mm). The first five volumes were discarded to allow for T1 equilibration effects.

We used FSL software (FMRIB’s Software Library, www.fmrib.ox.ac.uk/fsl) version five to analyse the functional data. Participants’ data (first-level analysis) were processed using the FMRI Expert Analysis Tool (FEAT). Functional images were realigned, motion corrected (MCFLIRT(Jenkinson *et al*, 2002)) and spatially smoothed with a 8mm full-width half-maximum Gaussian kernel. A high-pass filter was applied (120s cut-off). All images were registered to the whole-brain echo-planar image (EPI) (i.e., functional image with the whole-brain field of view), and then to the structural image of the corresponding participant (MPRAGE) and normalised to an MNI template, using linear registration with FSL FLIRT.

The five explanatory variables (EVs) that we used were the onset times of the standard, target, emotional, novel and neutral pictures. They were modelled as 1s events and convolved with a canonical double-gamma response function. We added a temporal derivative to the model to take into account possible variations in the haemodynamic response function. To capture residual movement-related artefacts, six covariates were used as regressors of no interest (three rigid-body translations and three rotations resulting from realignment). We used four contrasts: target-neutral, emotion-neutral, novel-neutral, and neutral-standard. However, in this study we only investigated the contrast of emotional-neutral. In the “second-level” analysis, we averaged the 3 blocks of the task for each participant using FEAT with Fixed Effects.

#### Region of interest analysis for all voxels within one cluster

For our main analysis, we pursued a region of interest (ROI) approach: For our salience type of interest – negative emotional salience – our primary hypothesis involved four regions that had been found to be most active in a former study using the same paradigm (Knolle *et al*, 2018). These four regions included the dopaminergic midbrain (ventral tegmental area/substantia nigra), the ventral and dorsal striatum, the bilateral hippocampus and the bilateral amygdala. The mask for the dopaminergic midbrain region was generated using the probabilistic atlas of Murty and colleagues (Murty *et al*, 2014) and has been used successfully in our own previous work (Ermakova *et al*, 2018; Knolle *et al*, 2018). Masks for all other regions were anatomically derived using the Harvard-Oxford subcortical structural atlas supplied with FSL. For planned group comparisons, we extracted contrast values (contrast of parameter estimates, or COPEs in FSL) for each individual from all the voxels within each of the four ROIs. We furthermore used the Featquery application in FSL to extract parameter estimates for individual event types within regions of interest for analysis presented in the supplementary materials. Average COPE values per region of interest were entered into a multivariate analysis of variance to compare groups.

#### Voxelwise permutation analysis

For analysis at the voxel-based resolution, all four ROIs were combined in one mask. For estimation of group comparison (higher level, or “third level”) statistics, we used permutation testing utilising the FSL randomise tool within our ROIs mask and the whole brain, with threshold-free-cluster enhancement, which enhances cluster-like structures but remains fundamentally a voxel-wise statistical testing method (Winkler *et al*, 2014). We used 5000 permutations and report significant results at p=0.05 or less following family-wise error correction for multiple comparisons, using the variance smoothing option (3mm) as recommended for experiments with small to modest sample sizes, as is common in fMRI research (Nichols and Holmes, 2002). We applied this method to our combined ROI and the whole brain. We used fslmeants to extract means from voxels revealing a significant group; we then used the extracted values purely for visualization of the group effect.

#### Resting cerebral blood flow

Interpretation of BOLD activation effects is complicated by difficulties in assessing whether any results are truly due to differences in evoked activation, or to baseline resting cerebral blood flow (CBF) (perfusion) differences “at rest” (Fleisher *et al*, 2009; Simon and Buxton, 2015). CBF could be altered by disease course or medication, as dopaminergic drugs act directly on the blood vessels and lead to vasodilation which increases CBF (Leenders *et al*, 1985). We therefore assessed resting CBF at baseline for all participants except for one PD patient without psychotic symptoms. For this assessment, we used a continuous arterial spin labelling (cASL) protocol described in Wang and colleagues (Wang *et al*, 2005) and adopted in other studies (Viviani *et al*, 2009). We used the following setup: relaxation time: 4000ms; echo time: 17ms; sequence: gradient-echo echo-planar imaging sequence with anterior-to-posterior phase encoding; multi-slice mode: interleaved; number of images: 120 with and without labelling; flip angle: 90°; in-plane resolution: 3.8×3.8×6mm; slice thickness: 6mm; matrix size: 64×64; field of view: 249×249mm; and bandwidth: 2442Hz/px. We inserted a 1s delay between labelling pulse and image acquisition. We used the SPM2 package (Wellcome Department of Cognitive Neurology, London; online at http://www.fil.ion.ucl.ac.uk) for realignment and stereotactic normalization to an EPI template (Montreal Neurological Institute, resampling size: 2 × 2 × 2 mm). Using the Perf_resconstruct_V02 SPM add-on software by Rao and Wang (Department of Radiology and Center for Functional Neuroimaging at University of Pennsylvania; online at http://www.cfn.upenn.edu/perfusion/software.htm), we reconstructed resting CBF values. We then used a ‘simple subtraction’ method (Wang *et al*, 2003). All volumes were smoothed using an isotropic Gaussian kernel of full width half-maximum (FWHM) of 8 mm prior to the analysis. We used the SPM PET basic models setup to generate our group statistics and then a one-way ANOVA with an explicit mask and an ANOVA normalisation. We also used the Marsbar toolbox to extract mean CBF for our regions of interest and then employed those values as covariates in our planned group comparisons for the task activations. Statistical analyses were generated using SPSS 21 (IBM Corp.).

#### Movement differences during fMRI scan

The task was split into 3 blocks of 11.5min. Blocks in which movement exceeded 3mm on average or 10mm maximum were excluded from the analysis. We only included participants with at least two blocks. We identified two PD patients without psychotic symptoms that had movement exclusion criterion in two out of three blocks and so they were excluded from all the analyses.

We compared the maximum and mean movement across the three blocks in two separate repeated measure ANOVAs (Table 2). We did not find any significant group, block or interactions effect, neither for mean movement nor for maximum movement (all p>0.1).

**Table 2.** Mean and maximum movement across testing blocks and groups.

|  | Group | Mean (SD) | Max (SD) |
| --- | --- | --- | --- |
| Block 1 | PD-Psychosis | 0.79 (0.63) | 2.22 (1.58) |
|  | PD+Psychosis | 0.74 (0.40) | 2.08 (1.31) |
|  | Controls | 0.46 (0.24) | 1.27 (0.82) |
| Block 2 | PD-Psychosis | 0.73 (0.63) | 2.21 (1.87) |
|  | PD+Psychosis | 0.80 (0.54) | 1.98 (1.24) |
|  | Controls | 0.54 (0.43) | 1.60 (1.45) |
| Block 3 | PD-Psychosis | 0.86 (0.64) | 2.70 (2.02) |
|  | PD+Psychosis | 0.68 (0.51) | 2.01 (1.48) |
|  | Controls | 0.68 (0.83) | 1.68 (1.56) |

PD-Psychosis: PD patients without psychosis, PD+Psychosis: PD patients with psychosis

## Results

### Behavioural results

Throughout the task, participants were asked to press a button in response to two target pictures - one for face stimuli and one for the scene stimuli. This ensured that participants maintained their attention throughout the task. In two separate repeated measure ANOVAs, we analysed the number of button presses and the reaction times in response to both target pictures together (Supplementary Figure 1 and 2, respectively). We found a significant effect for the number of misses across blocks (F(2)=3.82 p=.025, 68.3%power), but no group effect or interaction. Bonferroni corrected post hoc-tests revealed that participants missed marginally more button presses in the third compared to the second block (p=.059). On average, participants failed to press the button on 6 target trials per block (mean run1=5.78 (SD=2.4); mean run2=5.45 (SD=2.4); mean run3=6.4 (SD=3.1)). Furthermore, we found a significant effect for reaction time across blocks (F(2)=6.31 p=.003, 88.9%power), but no group effect or interaction. Bonferroni corrected post hoc-tests revealed that participants reacted significantly faster to target images in block 1 compared to block 2 (p=.014) and block 3 (p=.019). On average, participants required between 500 and 600 ms (mean run1=.531 (SD=.1); mean run2=.555 (SD=.1); mean run3=.560 (SD=.1)) to make a response, which is consistent with previous findings (Knolle *et al*, 2018).

**Figure 2.**
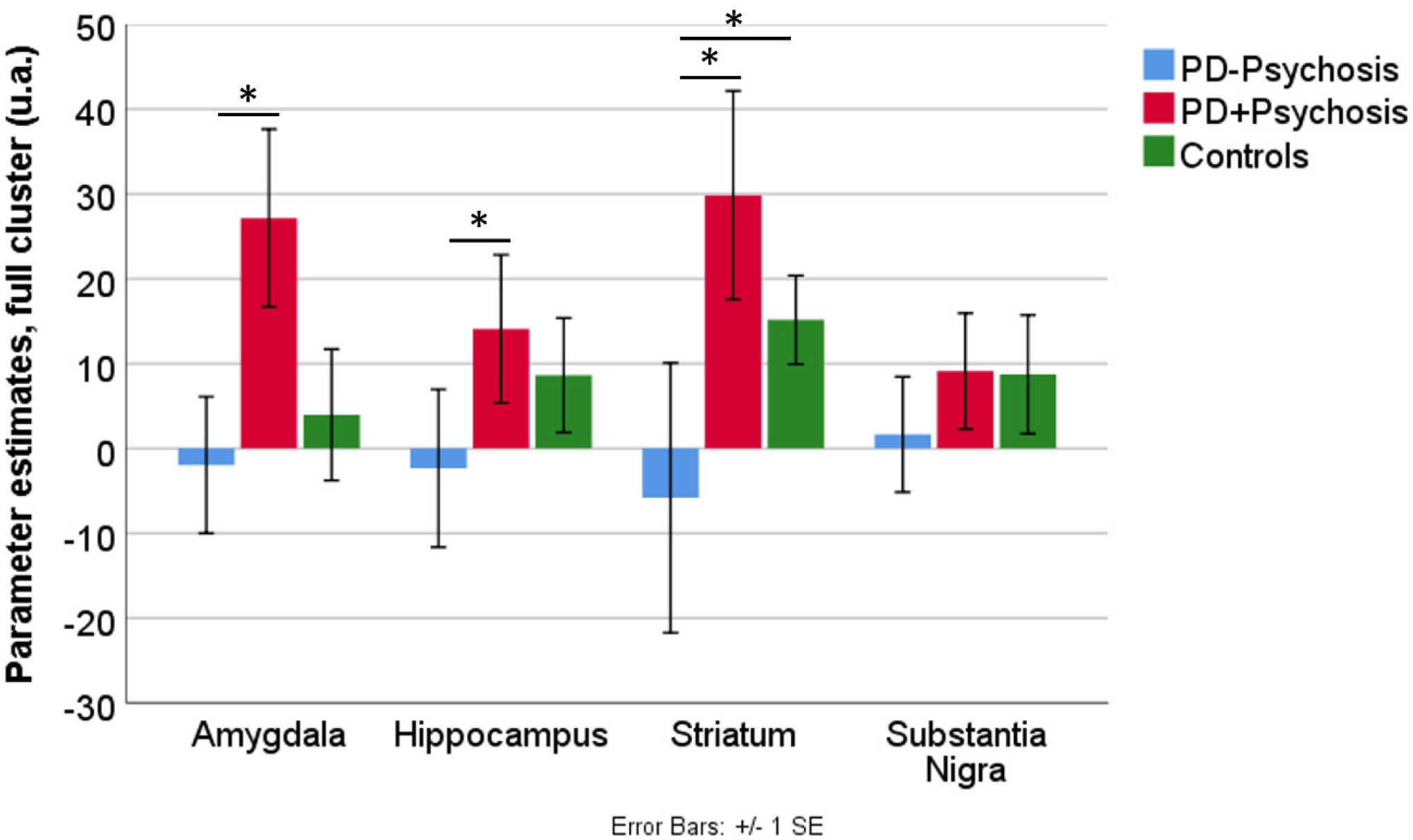
Bar chart shows mean contrast (COPEs, FSL) values extracted from all voxels of each region of interest and significant group effects (values uncorrected for covariates). Error bars show ±1 SE. *p<.05. PD-Psychosis: PD patients without psychosis, PD+Psychosis: PD patients with psychosis.

### Imaging results

#### Group analysis of resting cerebral blood flow

The one-way ANOVA on resting CBF data did not reveal any significant group differences.

#### fMRI activation to emotional salience

In our main analysis, we investigated potential group differences in emotional salience related activation, while controlling for resting CBF. We extracted the mean BOLD activation (COPE, contrast of parameter estimates between neutral and emotional oddballs) as well as the mean resting CBF from each individual region used in the ROI cluster. We conducted a multivariate analysis of variance to determine whether there were any statistically significant differences between the Parkinson’s patients with psychotic symptoms, Parkinson’s patients without psychotic symptoms and healthy controls on the BOLD activation per region controlling for CBF in the corresponding region (Figure 2).

The multivariate test revealed a significant group effect on brain activation in response to negative emotional salience within the ROIs, controlling for resting CBF in ROIs respectively, Pillai’s V=0.33, F(8,88)=2.14 p=.036. Tests of between-subjects effects furthermore revealed significant group effects in amygdala bilaterally, F(2,46)=4.62 p=.015, partial η2=.17, 75.3% power, the hippocampus bilaterally, F(2,46)=3.31 p=.045, partial η2=.13, 59.9% power, and the striatum, F(2,46)= 3.20 p=.050, partial η2=.12, 58.4% power. As a control analysis, we ran the multivariate analysis without controlling for CBF. These results (presented in the supplementary materials) are very similar and this supports the conclusion that they are not driven by differences in the CBF.

In the amygdala, we found bilaterally significantly greater (p=.004) activation in PD patients with psychotic symptoms (mean: 28.51, SE 9.8) compared to those without (mean: -8.82, SE 7.6). Controls (mean: 4.95, SE 9.2) did not significantly differ from either patient group.

In the hippocampus bilaterally we found significantly greater (p=.028) activation in PD patients with psychotic symptoms (mean: 14.84, SE 9.0) compared to those without (mean: - 10.85, SE 7.1). Controls (mean: 11.41, SE 8.5) had marginally significantly greater activity compared to PD patients without psychotic symptoms but this did not significantly differ from PD patients with psychotic symptoms.

In the striatum we found significantly greater (p=.017) activation in PD patients with psychotic symptoms (mean: 32.33, SE 14.7) compared to those without (mean: -13.37, SE 11.5). Controls (mean: 12.4, SE 13.9) did not significantly differ from either patient group.

#### Exploratory correlations of symptom scores, brain responses and medication

We conducted exploratory Pearson correlations between medication, symptom scores and brain activations.

Importantly, in PD patients with psychotic symptoms, we found a positive correlation between LED and the BOLD activation in the ROIs (bilateral amygdala: r=.696, p=.017, bilateral hippocampus: r=.624, p=.040, substantia nigra: r=.678, p=.022, striatum: r=.721, p=.012). Furthermore, LED was positively correlated to BDI score (r=.641, p=.034) and LED and apathy score (AES, r=.891, p=.007). There was also an additional negative correlation between LED and the GAF disability score (r=-.603, p=.050). In the same patients, we found, however, that the BDI score was positively correlated to resting CBF bilaterally in the hippocampus (r=.631, p=.015), the amygdala (marginally significant: r=.515, p=.059) as well as the substantia nigra (r=.646, p=.013). We did not find any such correlations in either of the other groups.

Pearson correlation analysis within each group did not reveal any significant interactions between BOLD activation and resting CBF. Furthermore, we did not find any significant correlations between symptom scores (GAF disability and BDI scores) and brain responses to negative emotional stimuli, except for one. In patients without psychotic symptoms, we found that the BDI score was positively correlated to resting CBF in the substantia nigra (r=.450, p=.036).

We used the Fisher r-to-z transformation to test whether the correlations in PD patients with psychotic symptoms were significantly different from the correlations in the other groups (see Supplementary Table 1). We found that the correlations of LED and BOLD activation as well as symptom scores were significantly different between the two patient group. Correlations between BDI and resting CBF in patients with psychotic symptoms differed significantly from those in controls, but not from the other patient group.

#### Voxelwise permutation analysis on negative emotional salience BOLD activation

In our ROI, we found two clusters within the striatum, the left putamen and left pallidum, in which groups significantly differed (Table 3, Figure 3 A, B).

**Table 3:** Significant activations from group analysis (GLM) on Emotional – Neutral Oddball (Negative emotional salience)

| Anatomical structure | Hemisphere | Cluster size<br>(voxel) | p-value | Peak Z score | Peak<br>coordinates |  |  |
| --- | --- | --- | --- | --- | --- | --- | --- |
|  |  |  |  |  | x | y | z |
| Region of interest, incl. bilateral amygdala, bilateral hippocampus, dorsal and ventral striatum, and dopaminergic midbrain |  |  |  |  |  |  |  |
| Putamen | L | 117 | 0.034 | 7.31 | -24 | 2 | 4 |
| Pallidum | L | 52 | 0.036 | 8.88 | -24 | -16 | 0 |
| Whole brain analysis |  |  |  |  |  |  |  |
| Parietal operculum/supramarginal gyrus | L | 189 | 0.043 | 10.98 | -52 | -26 | 24 |
| Planum temporale/parietal operculum | L | 60 | 0.044 | 11.91 | -48 | -40 | 18 |
| Central opercular cortex/precentral gyrus | L | 22 | 0.047 | 12.2 | -54 | 0 | 6 |
| Postcentral gyrus/supramarginal gyrus | L | 19 | 0.049 | 10.44 | -42 | -26 | 40 |

FSL randomise, 5000 permutation, FWE cluster-corrected results.

**Figure 3.**
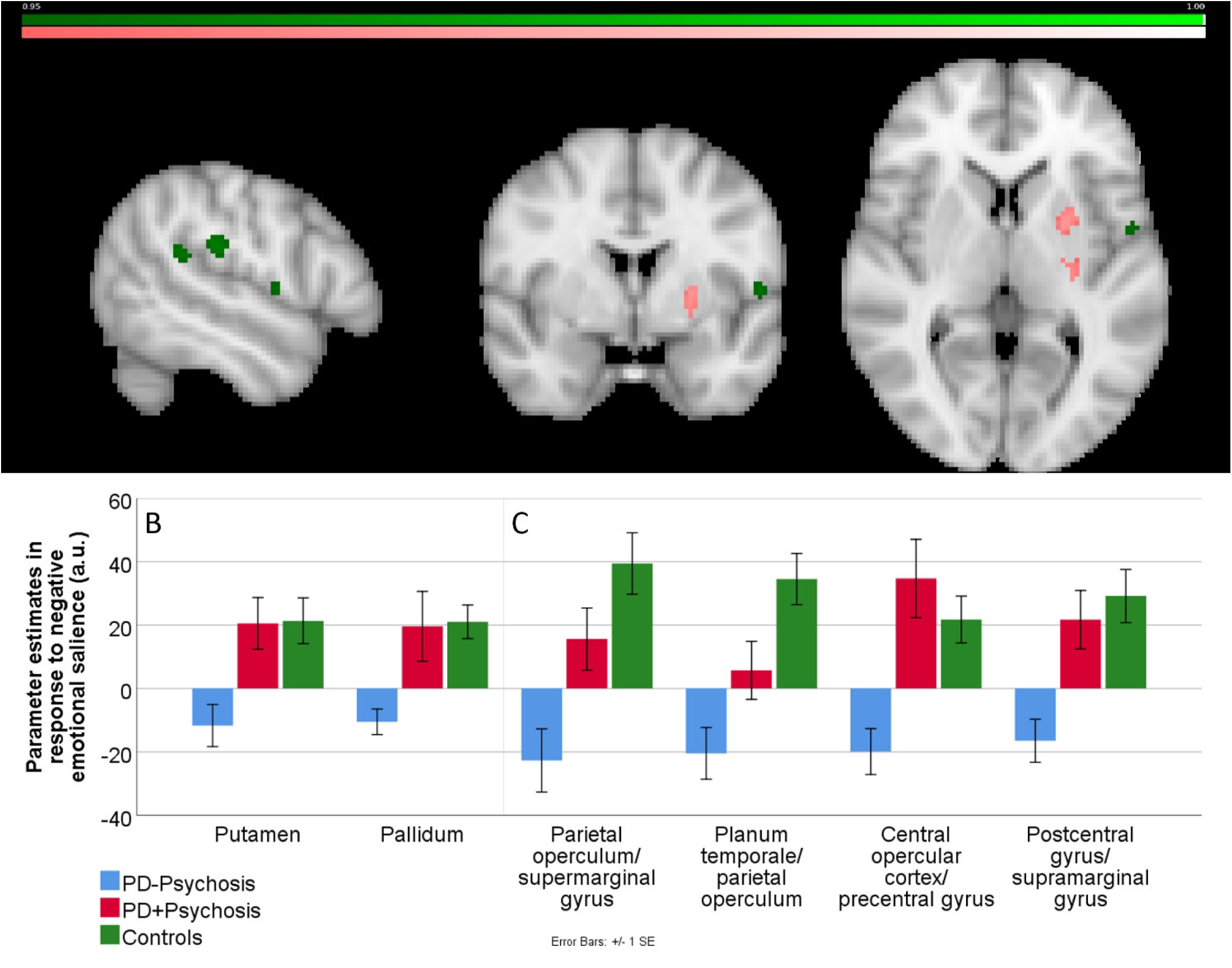
Group effects for the region of interests (ROI) as well as whole brain analysis showing activation associated with emotional salience processing (emotional oddballs versus neutral oddballs). A) Significant brain activations (slice location: x=-52, y=0, z=4) from the ROI analysis in putamen and pallidum (pink-to-white colour coding; maximal difference at x=0, y=-20, z=-6); and whole brain effects (dark green-to-light green colour coding) revealed four clusters including the pre- and postcentral gyrus and the supramarginal gyrus. Colour bars show significance level from p=0.05 (0.95) and lower (1). B/C) Bar charts showing extracted parameter estimates from significant clusters on the group level analysis as determined by the FSL randomised ANOVA, to visualise group differences. B) shows results of ROI analysis; C) results of whole brain analysis. Error bars show ±1 SE. PD-Psychosis: PD patients without psychosis, PD+Psychosis: PD patients with psychosis. L: left, R: right

Furthermore, a whole brain analysis revealed significant differences in the parietal operculum, the supramarginal gyrus, planum temporale and pre- and postcentral gyrus (Table 3, Figure 3 A, C).

We extracted parameter estimates from the significant clusters to visualise the group differences (Figure 3 B and C).

Additionally, we extracted the parameter estimates separately in response to emotional and neutral oddballs, as the conditions which define the contrast of interest. These are presented in the supplementary materials (Supplementary Figure 3) for all regions within the ROI. The parameter estimates indicate the potential drivers of the COPE (contrast of parameter estimates) effect.

## Discussion

In the current study we investigated negative emotional salience in PD patients with and without psychotic symptoms and compared them to healthy controls. Based on previous studies and the literature (Ermakova *et al*, 2018; Garofalo *et al*, 2017; Knolle *et al*, 2018), we hypothesised there would be altered brain activity in the striatum, substantia nigra, hippocampus and amygdala in both patients’ groups compared to control subjects, with an intermediate alteration in PD patients without psychotic symptoms. In line with our hypothesis, we found significant differences between both patient groups. PD patients with psychotic symptoms revealed strongly enhanced brain responses in the striatum, the hippocampus and the amygdala compared to PD patients without psychotic symptoms. PD patients with psychotic symptoms showed enhanced responses compared to controls but the difference did not reach significance. PD patients without psychotic symptoms only differed from controls in the striatum.

Our study is the first to investigate emotional salience processing in PD patients with and without psychotic symptoms. Our findings suggest that salience processing differentiates the two patient groups, and that overactivation within the hippocampal-striatal-midbrain circuits might contribute to the occurrence of psychotic symptoms in PD patients. Importantly, our study controls for putative dopaminergic medication effects on the baseline BOLD signal strength by assessing the resting cerebral blood flow (resting CBF) in all groups.

The current study reveals that PD patients with psychotic symptoms show a strongly enhanced response to salience in the striatum, amygdala and hippocampus compared to PD patients without psychotic symptoms. The pattern of activation in the PD patients with psychotic symptoms is reversed to that which has been reported in patients with primary psychosis (Knolle *et al*, 2018). Interestingly, however, in that work we also found that the stronger the psychotic symptoms the stronger the activation in response to emotional salience which is in line with the results in the PD patients with psychotic symptoms. Correspondingly, an exploratory analysis revealed that in PD patients with psychotic symptoms, the dose of dopaminergic medication as measured by LED, positively correlated with the BOLD activation in all ROIs (i.e., bilateral amygdala, bilateral hippocampus, substantia nigra). In addition, the dopaminergic medication dose was positively linked to measured depression (BDI) and apathy (AES), as well as negatively linked to global functioning (GAF disability). In patients without psychotic symptoms, we did not find any significant correlations between brain activation and psychopathology or medication, controls also not showing any correlations between brain activation and psychopathology. There was no significant difference in the overall medication dose between both patient groups. We therefore suggest a potential imbalance in the interaction between medication dependent tonic dopamine levels and phasic dopamine responses to sensory input in PD patients with psychotic symptoms. Our study indicates a link between the use of medication, processing alterations of salient stimuli as well as symptom scores. This is consistent with a study showing that the administration of a dopamine agonist (pramipexole or ropinirole) in young medication-naïve PD patients led to an increase in aberrant motivational salience by facilitating arbitrary and illusory associations between stimuli and rewards, and by faster reaction times to task-irrelevant stimuli as well as a slight increase in psychotic like symptoms (Nagy *et al*, 2012). Furthermore, in PD patients with psychotic symptoms only, we found positive correlations between the resting CBF within the four ROIs and depression severity, linking higher depression severity with higher resting CBF. When comparing the correlations across groups, PD patients with psychotic symptoms were only significantly different from the correlations in healthy controls. As the correlations were stronger in patients with, compared to without, psychotic symptoms, the results provide some additional indication for the mechanistic link between risk factors like depression and PD psychosis. Bold signal strength has been reported to depend on CBF levels (Simon and Buxton, 2015) but importantly we did not find that there were any significant differences in these parameters between groups. Therefore, it is unlikely that this correlation could fully explain the opposing signal between the two patient groups.

Our results are consistent with prior evidence that the use of dopaminergic medication is linked to the development of psychotic symptoms in PD patients (Zahodne and Fernandez, 2008). However, we still lack a full mechanistic explanation of why the use of dopaminergic drugs lead to psychotic symptoms in some patients but not in others. The aberrant salience hypothesis of psychosis suggests, first, that a dysregulated dopaminergic system in the mesolimbic system leads to the attribution of salience to otherwise irrelevant signals (Kapur, 2003); and, second, that these irrelevant signals are taken as valid information, and integrated by seemingly plausible top-down explanations, which supports the development of delusions and hallucinations.

With regard to the first prerequisite, PD patients show a clear dopaminergic pathology, which may involve dysregulation in some patients. Deficits in critical reasoning and accepting hasty cognitive explanations have often been reported in psychosis, mainly in schizophrenia but also other psychotic disorders (Garety *et al*, 2005; Lincoln *et al*, 2010). ‘Jumping to conclusions’ reflects a bias in critical reasoning where individuals draw a conclusion based on too little information for making an informed decision. In psychosis, ‘jumping to conclusions’ is considered a trait contributing to developing delusions (Garety and Freeman, 2013), as individuals who jump to conclusions might be prone to accepting implausible ideas and disregard alternative explanations. Djamshidan and colleagues were able to detect a bias in generating and accepting abnormal explanations for aberrantly salient stimuli in medicated (Djamshidian *et al*, 2012) and unmedicated (de Rezende Costa *et al*, 2016) PD patients. We speculate that this could relate to a cortical pathology, which is now well recognised in Parkinson’s disease (Kövari *et al*, 2003; Mattila *et al*, 2000). In our current study, we report some exploratory ‘jumping to conclusions’ results on our cohort of patients and controls in the supplementary materials given that only some of the patients/controls completed this part of our study. The results indicated that PD patients with psychotic symptoms showed a ‘jumping to conclusion’ bias by sampling less information before making a decision, which therefore also had a lower probability of being correct. This bias was not present in PD patients without psychotic symptoms or controls. We therefore suggest that the development of psychotic symptoms in PD patients may result from a combination of aberrantly enhanced salience signals in the striatal-hippocampal-midbrain circuits and deficient cognitive reasoning (possibly cortical) processes.

In conclusion, our study provides evidence for the first time that aberrant striatal, hippocampal and amygdala signalling during processing of non-motivational salient stimuli differentiates PD patients with and without psychotic symptoms. Furthermore, the results indicated impaired top-down processing in PD patients with psychotic symptoms. Taken together, the results suggest that enhanced signalling in these regions and deficient top-down cognitive regulations, possibly lead to the development of psychotic symptoms, in a similar way as that proposed in the ‘aberrant salience hypothesis’ of psychosis.

## Data Availability

Data available on request.

## Acknowledgements

We would like to thank Lisa Ronan and Claire O’Callaghan for fruitful discussions and comments.

